# Dynamic Functional Connectivity States in Narcolepsy Type 1: Distinct Patterns from Acute Sleep Deprivation and Associations with Clinical Measures of Sleepiness

**DOI:** 10.1101/2025.08.06.25333088

**Authors:** Wenwei Zhu, Fulong Xiao, Mengmeng Wang, Xiaosong Dong, Fang Han, Ning Ma

## Abstract

**Background and Objectives:** Narcolepsy Type 1 (NT1) is a neurological disorder caused by hypocretin deficiency, leading to excessive daytime sleepiness and cataplexy. This study characterized dynamic functional connectivity (dFC) states in NT1 patients, acute sleep-deprived (SD) individuals, and healthy controls, and explored how these states relate to clinical measures of sleepiness and arousal.

**Methods:** In this study, resting-state co-fluctuation analysis was employed to identify recurring brain states and compare group differences in state dwell time, transition probabilities, and interaction strength. Associations between dFC properties and clinical metrics (Epworth Sleepiness Scale [ESS] scores, mean sleep latency from MSLT) were also investigated.

**Results:** Five distinct resting-state co-fluctuation states were identified. NT1 patients showed significantly longer mean dwell time and higher fraction rate in State 3, characterized by synchronized activity between the salience/ventral attention network (SN/VAN) and sensorimotor network (SMN) with antagonistic co-fluctuations to the visual network (VIS), compared to both SD and control groups. They also exhibited increased reciprocal transition probabilities between State 3 and State 5. Group-specific differences in co-fluctuation strength were observed across multiple states, with NT1 showing distinct alterations in interactions involving the striatum, limbic system, and attentional networks. Moreover, the fraction rate of State 5 negatively correlated with ESS scores, while the fraction rate of State 3 negatively correlated with mean sleep latency from MSLT in NT1 patients, indicating that increased occupancy of certain states was associated with less subjective sleepiness and greater arousal instability.

**Conclusion:** These findings highlight the role of chronic hypocretin-mediated arousal failure versus acute homeostatic sleep pressure in shaping network co-fluctuation patterns, characterized by thalamocortical disconnection, cortical dysregulation, and enhanced striatal-limbic connectivity. This state might be specific to hypocretin deficiency and suggests that dFC states may serve as potential biomarkers for sleep-wake disorders.

## Introduction

Narcolepsy with cataplexy (Narcolepsy Type 1, NT1) is a chronic neurological disease characterized by sleep-wake dysregulation, including excessive daytime sleepiness (EDS), sleep paralysis, cataplexy, and disturbed sleep.^1–4^ The loss of hypocretin (orexin) is the core pathological mechanism underlying NT1.^5^ This neural dysfunction disrupts arousal regulation,^6^ leading to instability in brain networks critical for maintaining wakefulness, such as the salience network, ventral attention network, and sensorimotor network.^7–10^ While static functional connectivity studies have identified aberrant interactions in these networks, the dynamic temporal organization of brain states, including how networks co-fluctuate and transition over time, remains poorly understood in NT1.

The absence of hypocretin disrupts neurotransmitter homeostasis,^11^ and may also affect brain activity. Neuroimaging studies in NT1 have revealed that hypocretin deficiency is associated with reduced metabolic activity in the thalamus and striatum.^12, 13^ Additionally, altered connectivity has been observed in the salience network and subcortical network, which are involved in attention and arousal regulation.^14^ These findings suggest that the loss of hypocretin impairs subcortical function and disrupts the integration of cortical and subcortical. However, comparing patients with NT1 with healthy controls may conflate two distinct mechanisms: irreversible brain activity changes due to hypocretin deficiency and reversible changes induced by sleepiness. Sleepiness is defined as a state of increased sleep propensity with impaired wake maintenance,^15^ manifesting in both patients with NT1and acute sleep-deprived healthy individuals. While both groups experience sleepiness,^6,16^ only patients with NT1 have irreversible hypocretin deficits. Acute sleep deprivation, a model of transient wakefulness impairment, provides a valuable comparator to NT1: while NT1 involves chronic hypocretin-mediated arousal failure, SD induces homeostatic sleep pressure without structural neural loss. Comparing patients with NT1, healthy controls, and individuals experiencing acute sleep deprivation can help disentangle chronic pathological arousal instability (NT1) from acute functional arousal deficits (SD), thereby identifying state-specific signatures of hypocretin-dependent versus sleep homeostatic dysfunction. Given that prior research exploring the neural basis of NT1 has not fully controlled the influence of sleepiness, the present study included an SD group to elucidate the endogenous pathological mechanism (the effects of hypocretin deficiency on brain function) of NT1.

Among numerous neuroimaging modalities, resting-state functional connectivity (rs-FC) offers unique advantages in revealing spontaneous interactions within brain networks. The typical symptom of NT1 (daytime sleep attacks) results from spontaneous sleep-wake dysregulation rather than being triggered by external tasks or stimuli.^3^ The transient and episodic nature of sleep attacks reflects instability in brain activity, which limits the effectiveness of static functional connectivity in investigating the neural mechanisms of NT1. Specifically, patients with NT1 often experience sleep attacks during which their brain suddenly enters a “sleep mode”.^17^ Sleep attacks occur over brief periods, with a rapid reorganization of brain network activity; however, static functional connectivity analysis fails to capture these transient network changes over time. In contrast, dynamic functional connectivity (dFC) has emerged as a powerful tool for capturing transient network changes by monitoring real-time functional interactions within the brain.^18^Given that current research has inadequately addressed real-time network reorganization during spontaneous sleep attacks, the analysis of dFC during resting state to assess temporal properties and dynamic state strengths may help uncover brain function reorganization in specific states and changes of spontaneous activity patterns associated with clinical relevance.

Key clinical metrics in NT1, including the Epworth Sleepiness Scale (ESS) and mean sleep latency from MSLT, reflect subjective and objective measures of sleepiness. We hypothesized that NT1 would exhibit distinct dynamic brain state properties compared to both healthy controls and acute sleep-deprived individuals, particularly in networks mediating arousal (SN/VAN, SMN) and sleep regulation (DMN, visual network [VIS]). In contrast, acute sleep-deprived individuals might exhibit transient alterations in state transitions or co-fluctuation strength, driven by homeostatic sleep pressure rather than structural neural dysfunction. Here, we employed resting-state co-fluctuation analysis to identify recurring brain states and characterize group differences in state dwell time, transition probabilities, and interaction strength among patients with NT1, healthy controls, and acute sleep-deprived participants. By integrating clinical measures (ESS, mean sleep latency from MSLT) with dynamic connectivity properties, this study aims to: 1) define distinct dynamic brain state profiles in NT1 and SD; 2) identify hypocretin-dependent vs. homeostatic sleep pressure-related alterations in network co-fluctuation; and 3) establish whether state dynamics correlate with clinical markers of sleepiness. These insights may uncover novel neural signatures of NT1 pathophysiology and inform targeted interventions for sleep-wake disorders involving dynamic brain state dysfunction.

## Methods

### Participants

This study recruited 52 drug-naïve adult patients diagnosed with NT1 according to the International Classification of Sleep Disorders-3 (ICSD-3, The diagnosis details in Supplemental Methods)^19^ and concentration of hypocretin-1 in cerebrospinal fluid (CSF) from the Sleep Medicine Center in Peking University People’s Hospital, as well as 50 healthy control participants. In addition, 43 healthy participants who experienced 24-hour SD were recruited from the Center for Sleep Research at South China Normal University. All participants were controlled for sleep quality, daytime sleepiness, circadian rhythms, and neuropsychiatric history. Detailed questionnaires and exclusion criteria are provided in Supplemental Methods. All patients with NT1 were first-time visitors and had never been prescribed psychiatric stimulants. The definitive diagnosis was subsequently confirmed through a nocturnal polysomnography followed by a multiple sleep latency test (MSLT). To quantify the severity of EDS across all participants (including NT1, healthy controls, and the SD group), the ESS was used. The ESS is an eight-item self-report questionnaire in which participants rate the possibility of falling asleep in various situations on a scale from zero to three. A cumulative ESS score of ten or higher indicates the presence of EDS.^20^ The exclusion criteria for both patients with NT1 and the two healthy groups are provided in Supplemental Methods.

### Polysomnography and MSLT Evaluation

Polysomnography and the MSLT were recorded only in patients with NT1 to a definitive diagnosis. A full nocturnal polysomnography was recorded on a Respironics LE-Series Physiological Monitoring System (Alice 6 LE, Philips, Murrysville, FL) from 10:00 p.m. to 6:00 a.m., followed by an MSLT the next day. Subsequently, we calculated the mean sleep latency across the five naps in the MSLT. Detailed information about PSG and MSLT is provided in Supplemental Methods.

### Experimental Protocol of Acute Sleep Deprivation

During the SD session, participants arrived at the laboratory at 7:00 p.m. and remained continuously awake throughout the night under the supervision of trained personnel, resulting in approximately 24 hours of total sleep deprivation. All participants were in good physical and mental health, and we also controlled for circadian rhythms and prior sleep debt. Details of the procedures are available in Supplemental Methods.

### Imaging Data Acquisition

Resting-state functional MRI (rs-fMRI) and high-resolution T1-weighted structural images were obtained using a 3T scanner (Siemens, Skyra, Germany) for both patients with NT1 and healthy controls. Patients with NT1 performed the MRI scan following the MSLT completion, while SD participants were scanned the morning after the SD protocol. Both resting fMRI scans and high-resolution T1-weighted structural images of SD participants were obtained using a 3T scanner (Siemens, Erlangen, Germany). All participants were instructed to resist sleeping, remain fully awake, refrain from moving, and keep their eyes open throughout the MRI scan. Additionally, emotional triggers were avoided during the scanning procedure to prevent cataplexy attacks. Details of MRI data acquisition parameters are described in Supplemental Methods.

### Preprocessing of Functional Imaging Data

Functional MRI data preprocessing was performed using the Data Processing and Analysis for Resting-State Brain Imaging V2.1 (DPABI V2.1), which works with Statistical Parametric Mapping (SPM8) implemented in the MATLAB platform (The MathWorks Inc., Natick, MA).^21^ The preprocessing of fMRI data included correction for slice timing and head motion, registration to 3D T1-structural images, normalization to the Montreal Neurological Institute template, and smoothing, as well as removal of linear trends. Finally, the resulting time series were projected on the cortical and subcortical surface of each participant, divided into 234 ROIs (200 cortical plus 34 subcortical). These areas are taken from the multi-resolution functional connectivity-based cortical parcellations developed by Schaefer and colleagues,^22^ including eight additional subcortical regions (bilateral caudate, putamen, pallidum, thalamus) and cerebellar parcels (26 cerebellum regions) from the Automated Anatomical Labeling atlas.^23^ Details of preprocessing parameters are described in Supplemental Methods.

### Dynamic Functional Connectivity

#### Edge (Co-fluctuation) Time Series

Edge-centric is a method for estimating time-varying function connections.^24^ This method precisely decomposes FC into its framewise contributions, providing a frame-by-frame account of interregional co-fluctuations over time, which we refer to as co-fluctuation or edge time series (ETS). A key advantage of this method is that ETS are estimated without specifying parameters or windowing, thereby avoiding many of the limitations associated with sliding window methods. ETS has been applied to study individual differences and the origins of brain systems. The anatomical underpinnings have been examined using in silico models. Further details on the computation of ETS can be found in Supplemental Methods.

#### ComBat Correction for Data Batch Effects

In neuroimaging studies, data collected from different machines or scanning sessions may introduce batch effects, which can potentially confound the analysis of brain networks. To eliminate these sources of variability, we applied the ComBat correction, a widely used statistical method for batch effect correction.^25^ Researchers initially applied ComBat to genomic datasets and subsequently adapted the method for neuroimaging.^26^ Details of ComBat are described in Supplemental Methods.

We used ComBat correction to integrate data from the two scanning sessions, reducing scanner-related variability and discrepancies in MRI acquisition parameters. This step enabled a direct comparison of dynamic functional connectivity across groups without the confounding of batch effects.

#### Clustering Analysis of ETS

We applied a k-means clustering algorithm on ETS to identify recurring functional connectivity patterns (states), as expressed by the frequency and structure of these states. The optimal number of clusters (K = 5) was determined using the elbow criterion applied to a cluster index computed from running k-means on all participants’ ETS data, with the number of clusters ranging from two to ten. The cluster index was defined as the ratio of the within-cluster sum distances to the between-cluster sum distances. Clustering results (clusters and output metrics) are reported for the optimal number of clusters. The five clusters, referred to as functional network connection states, describe five connectivity patterns that participants move through over time. The total number of dFCs and participants per state was also recorded. It is important to note that not all individuals exhibited dFC in every state. Therefore, the number of participants in each state is a subset of the entire participant cohort. Additional clustering details are described in Supplemental Methods.

#### Dynamic Network connectivity

We assigned 234 ROIs to 19 networks to construct network connectivity based on the pre-defined cortical parcellation and AAL template.^22, 23^ The 19 networks included executive control networks (CON: ContA, ContB, and ContC), default mode networks (DMN: Default A, Default B, and Default C), dorsal attention networks (DAN: DorsAttnA and DorsAttnB), limbic network (Limbic), salience/ventral attention networks (SN/VAN: Sal/VentAttnA and Sal/VentAttnB), somatomotor networks (SMN: SomMotA and SomMotB), temporoparietal network (TempPar), visual networks (VIS: VisCent and VisPeri), subcortical networks (SUB: striatum, thalamus), and cerebellar network (Cerebellum). Finally, intra-network and inter-network connectivity within each time point was calculated among the 19 networks for further analysis.

### Statistical Analysis

#### Group Differences in Dynamic Connectivity: Temporal Properties and Strength

The four temporal properties metrics that provide clustering measures are mean dwell time (average time participants spend in each state before changing to another), fraction rate (percentage of total time participants spend in each state), transition probability (the probability of transitioning between states), and number of transitions (the number of transitions between states and represents the reliability of each state). Group differences in dwell time, fraction time, transition probability, and number of transitions between the control group (Control), SD group, and NT1 were examined using ANOVA (*p*< 0.05, Bonferroni correction) with BMI as a covariate. Subject-specific medians corresponding to each group-level state were estimated, and ANOVA was used to compare the co-fluctuation strength of each state at each unique regional pairing (total of 171 pairings; *p*< 0.05, Bonferroni correction) between three groups with BMI as a covariate.

#### Demographic and Clinical Data Analysis

Statistical analyses were performed using SPSS Statistic,^27^ release version 24.0 (Chicago, IL, USA). Group differences in age, BMI, and ESS scores were compared using ANOVA. Pearson’s chi-square test was applied to compare categorical variables. We conducted Pearson’s correlation analyses between temporal properties and clinical data (ESS scores and mean sleep latency from MSLT) in the NT1 group, controlling for BMI as a covariate. The statistical significance threshold was set at *p*< 0.05 and corrected by Bonferroni correction.

### Standard Protocol Approvals, Registrations, and Patient Consents

The present study was approved by the Medical Ethics Committee of Peking University People’s University and the Medical Ethics Committee of South China Normal University. Written informed consent was obtained from all participants after a full explanation of the procedure involved. The research was conducted in accordance with the Declaration of Helsinki (2002 version).

## Results

### Demographic

We excluded five participants because of excessive head motion (2 participants from NT1 group and 3 participants from SD group), leaving 50 patients with NT1 (36 % female; mean ± SD = 24.36 ± 3.97 years), 40 healthy participants who experienced acute sleep deprivation (42.5 % female; mean ± SD = 24.85 ± 2.08 years), and 50 healthy controls (50 % female; mean ± SD = 23.76 ± 3.46 years). Table 1 summarizes demographic information, ESS scores, and mean sleep latency from MSLT across the three groups, along with group comparisons. The study included 50 patients with NT1, 50 healthy control participants, and 40 healthy SD participants. Significant group differences were observed in both ESS scores and BMI. Patients with NT1 had the highest ESS scores, followed by the SD group, with the control group scoring the lowest. A similar pattern was observed for BMI, which was highest in patients with NT1, followed by the control group, and lowest in the SD group. No significant group differences were found in age and gender variables.

**Table 1.** Participants’ characteristics including demographics, polysomnography measures, and questionnaire scores.

|  | NT1 (n = 50) | Control (n = 50) | SD (n = 40) | P value | Post-Hoc |
| --- | --- | --- | --- | --- | --- |
| **Demographics** |  |  |  |  |  |
| Male/female | 18/32 | 25/25 | 17/23 | 0.373 | - |
| Age (years) | 24.36 (3.97) | 23.76 (3.46) | 24.85 (2.08) | 0.311 | - |
| BMI (kg/m<sup>2</sup>) | 26.74 (4.44) | 24.17 (1.36) | 20.15 (2.01) | < 0.001 | NT1 > SD***; NT1 > Control ***; Control > SD*** |
| Disease onset-age (years) | 12.18 (4.2) | - | - | - | - |
| Cataplexy | 50/50 | - | - | - | - |
| **Epworth Sleepiness Scale** | 18.04 (3.89) | 4.17 (1.47) | 11.55 (2.51) | < 0.001 | NT1 > SD***; NT1 > Control ***; SD > Control *** |
| **Multiple Sleep Latency Test** |  |  |  |  |  |
| Latency at MSLT (mins) | 1.76 (1.51) | - | - | - | - |
| Number of SOREMPs at MSLT | 2.5 (1.9) | - | - | - | - |
| **Polysomnography** |  |  |  |  |  |
| TST (mins) | 443 (112.3) | - | - | - | - |
| Latency at PSG (mins) | 7.2 (3.9) | - | - | - | - |
| Sleep efficiency (%) | 88.5 (21.4) | - | - | - | - |
| WASO (mins) | 37.1 (25.6) | - | - | - | - |
| REM (mins) | 112.77 (46.3) | - | - | - | - |
| AHI | 4.29 (3.86) | - | - | - | - |
| **Biomarker** |  |  |  |  |  |
| HLA DQB1*06:02 positive (%) | 100% | - | - | - | - |
| Hypocretin level in cerebral spinal fluid (pg/mL) | 26±19.7 (n = 6) | - | - | - | - |

**Note:** Values are given as mean (Standard Deviation). NT1 = Narcolepsy with cataplexy; SD = Acute Sleep Deprivation; ESS = Epworth Sleepiness Scale; IH: Idiopathic Hypersomnia; MSLT: Multiple Sleep Latency Test, TST: Total Sleep Time, WASO: Wake After Sleep Onset, AHI: Apnea-hypopnea Index; BMI = Body Mass Index. ANOVA was used to compare group differences. Chi-square test was used for gender difference. \*\*\* $p < 0.001$ .

### Dynamic Co-fluctuation State Analysis

#### Temporal Properties

We identified five recurring patterns of resting-state co-fluctuation that were consistently observed within and across participants. The cluster centroids of the five states, along with their overall occurrence rates, are shown in Figure 1A. States 1 through 4 were relatively infrequent (around 6% of the total occurrences in State 1, 8% in State 2, 7% in State 3, and 5% in State 4) but exhibited stronger co-fluctuations compared to the more prevalent State 5. Figure 1B shows the top 10% of co-fluctuation edges across these states.

**Figure 1.**
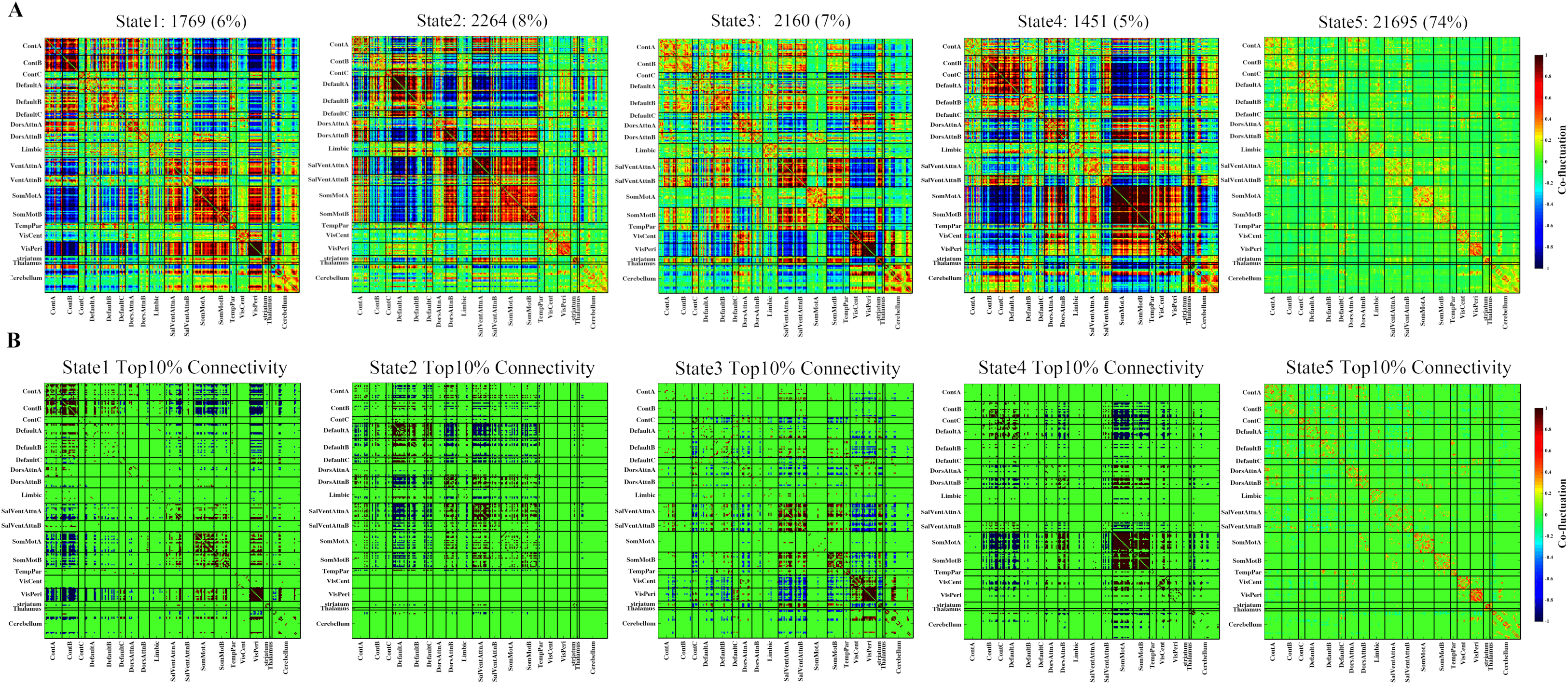
Results of the clustering analysis per state. **(A)** Cluster centroids for each state. The total number of occurrences and the percentage of total occurrences are listed above each cluster median. **(B)** Only 10% of the functional connectivity matrix in each state is shown, representing the strongest connections.

State 1 (SN/VAN-VIS-SMN synchronized state with CON antagonism) was characterized by intra- and inter-network co-fluctuations among the SN/VAN, SMN, and VIS, which were collectively anticorrelated with CON. State 2 (SN/VAN-SMN synchronized state with DMN asynchrony) is characterized by intra- and inter-network synchronization between SN/VAN and SMN, with anticorrelated interactions with DMN. State 3 (SN/VAN-SMN synchronized state with VIS antagonism) also involved synchronization between SN/VAN and SMN, but with anticorrelation with VIS. State 4 (SMN-DAN synchronized state with CON and thalamus antagonism) was characterized by co-fluctuations between SMN and the DAN, accompanied by anticorrelation with both CON and the thalamus. State 5 showed sparse, largely within-network co-fluctuations, reflecting a baseline pattern of reduced network interaction. Group-specific cluster centroids of NT1, health controls, and ASD closely resembled the centroids derived from the full-sample k-means clustering (Supplemental Figure 1).

As shown in Figure 2A, significant group differences were observed in the mean dwell time of State 2 and State 3. The NT1 group showed a significantly longer mean dwell time in State 3 compared to both the control and SD groups, while the SD group showed a shorter mean dwell time in State 2. Similar group differences were observed in the fraction rate across states (Figure 2B). The NT1 group exhibited the highest fraction rate in State 3 compared to both the SD and control groups. In contrast, the control group exhibited the highest fraction rate in State 2, while the SD group showed the highest fraction rate in State 1. No significant group differences were found in the fraction rate or mean dwell time of State 4 and State 5. Detailed statistics information is provided in Supplemental Table 1 and Supplemental Table 2.

**Figure 2.**
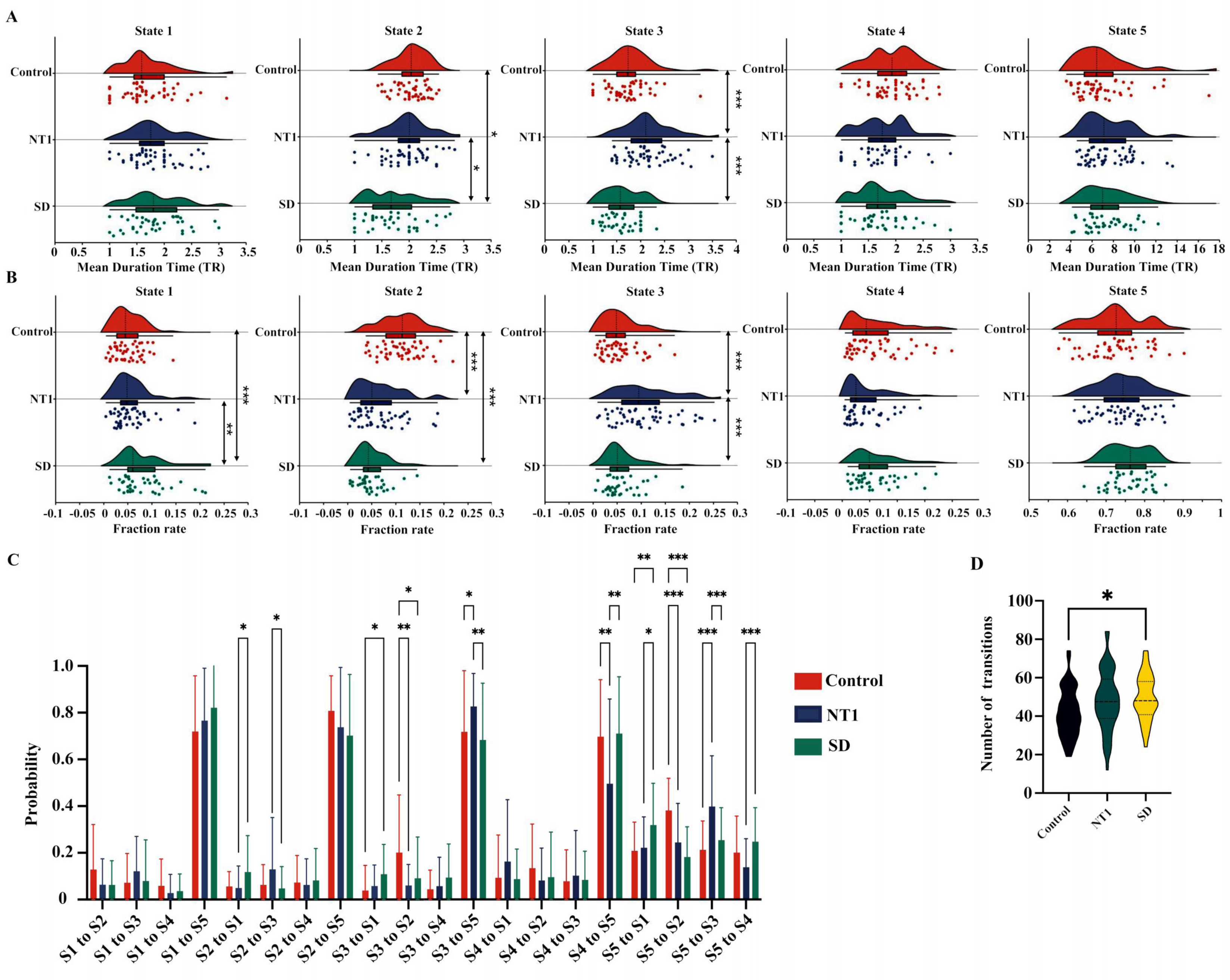
Results of temporal properties of functional connectivity state analysis. **(A)** Mean dwell time (i.e. number of consecutive windows spent in each state before switching), **(B)** the fractional rate of total subjects spent in each state as measured by percentage, **(C)** the probability of state transition in each group and **(D)** number of transitions (i.e. switching between states) is depicted for the Narcolepsy with cataplexy (NT1), Acute Sleep Deprivation (SD) group and control group (Control). Asterisks indicate a significant group difference (* *p*< 0.05; ** *p*< 0.01; *** *p*< 0.001, Bonferroni corrected).

We further examined transition probabilities between the five states (Figure 2C, detailed statistics in Supplemental Table 3). The NT1 group exhibited significantly higher probabilities of transitioning between State 3 and State 5 reciprocally, compared to both the control and SD groups. They also exhibited a higher probability of transitioning from State 2 to State 3 than the SD group, but were less likely to transition from State 4 to State 5 than both other groups. The SD group, by contrast, demonstrated a stronger tendency to transition from State 5 to State 1 compared to both the NT1 and control groups, and was more likely than NT1 patients to transition from State 2 or State 3 to State 1, as well as from State 5 to State 4. No significant differences were observed between the NT1 and the control group for these transitions. Additionally, the control group was more likely than both other groups to transition from State 5 to State 2.

As shown in Figure 2D, the control group exhibited significantly fewer transitions between brain states compared to the SD group. There was also a trend toward fewer transitions in the control group compared to the NT1 group (NT1: *M* ± *SD* = 48.18 ± 15.66; Control: *M* ± *SD* = 42.02 ± 12.61; SD: *M* ± *SD* = 49.53 ± 11.88; F (2, 137) = 4.08; *p* = 0.019; post hoc result: Control vs NT1, *p* = 0.075; Control vs SD, *p* = 0.031; NT1 vs SD, *p* = 0.46; Bonferroni corrected).

Overall, these findings suggest that patients with NT1 spent more time in State 3, which was characterized by synchronized activity between SN/VAN and SMN, while exhibiting antagonistic co-fluctuations with VIS. Moreover, patients with NT1 showed a higher probability of transitioning between State 3 and State 5 reciprocally, indicating an increased tendency to shift into and remain in this state.

#### Strength of Dynamics States

We compared co-fluctuation strength across NT1, control, and SD groups for each state to identify network interactions that might reflect pathological features specific to NT1. Co-fluctuations that significantly differed in NT1 compared to both the control and SD groups were considered potential pathological features of NT1. Figure 3 summarizes the group differences in co-fluctuation strength across states and Figure 4 summarizes potential pathological features of NT1. In State 1, six network pairings were distinct in NT1: higher co-fluctuations were observed between SalVentAttnA and ContA, SalVentAttnB and DefaultB, and the Cerebellum and Limbic, while lower co-fluctuation was found between SalVentAttnA and SalVentAttnB, SalVentAttnB and SomMotB, and SalVentAttnB and thalamus. In State 2, five network pairs showed NT1-specific differences: co-fluctuation between SalVentAttnA and DefaultA, Striatum and DefaultA, Striatum and DefaultB, and Striatum and Limbic were significantly increased in NT1, while co-fluctuation between SalVentAttnA and Striatum was decreased. State 3 revealed three NT1-specific alterations: higher co-fluctuation between the Striatum and Limbic, and between VisCent and DorsAttnA, whereas lower co-fluctuation was observed between VisCent and DefaultB. No significant group differences were found in State 4. In State 5, NT1 patients showed increased co-fluctuation between ContC and ContB, Striatum and Limbic, and Cerebellum and Limbic, while exhibiting decreased co-fluctuation between DefaultC and ContA, VisCent and DefaultB, and Cerebellum and DorsAttnB. Detailed statistical results for each state are provided in Supplemental Table4 to Supplemental Table 7.

**Figure 3.**
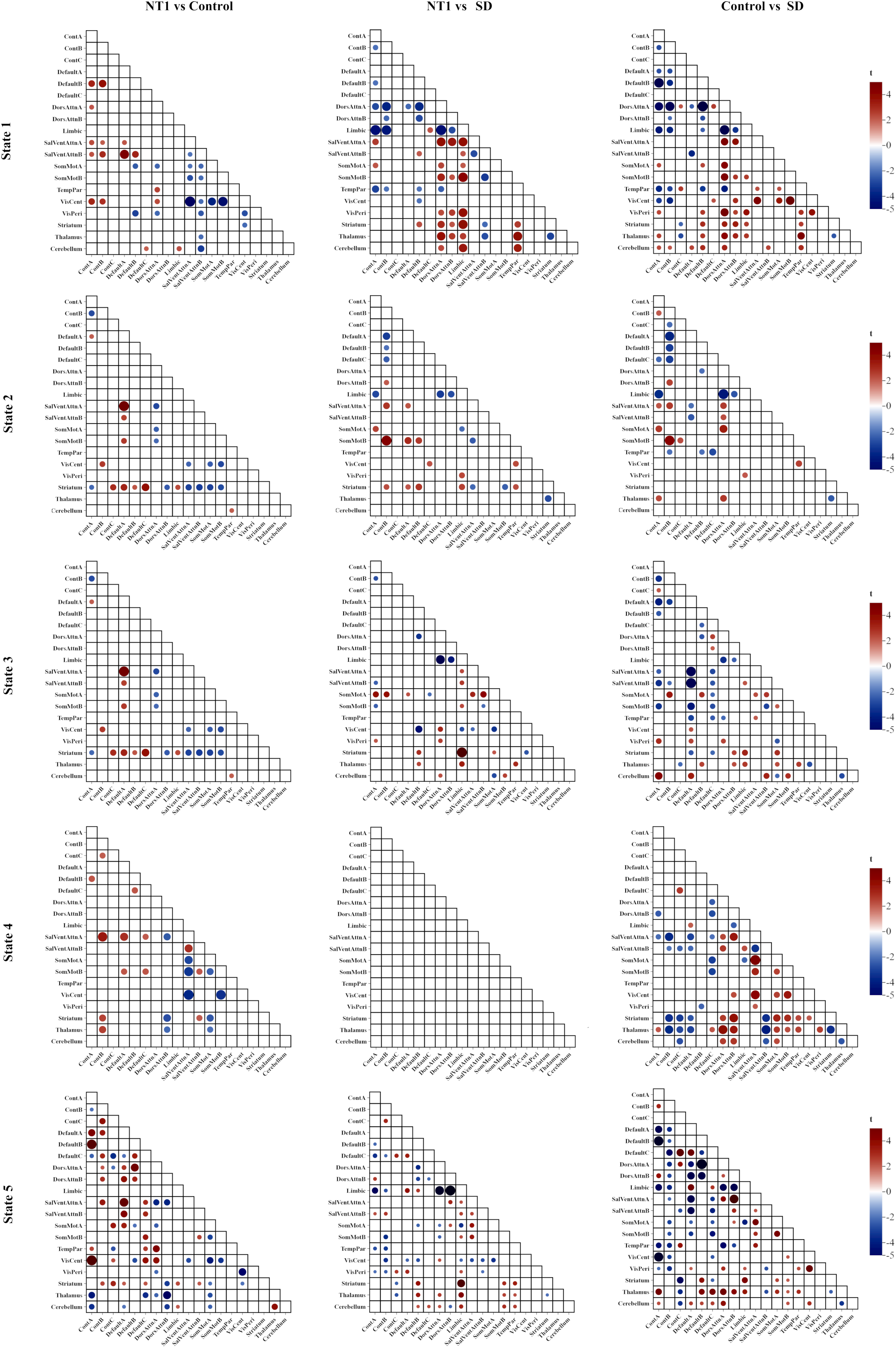
Functional connectivity state results. Functional connectivity in each state, where narcolepsy with cataplexy had a weaker or stronger co-fluctuation pattern in comparison to the control and SD (acute sleep-deprivation) groups. NT1 vs Con refers to the results compared to narcolepsy with cataplexy and the control group; NT1 vs SD refers to the results compared to narcolepsy with cataplexy and the acute sleep deprivation group; Con vs SD refers to the results compared between the control group and the acute sleep deprivation group.

**Figure 4.**
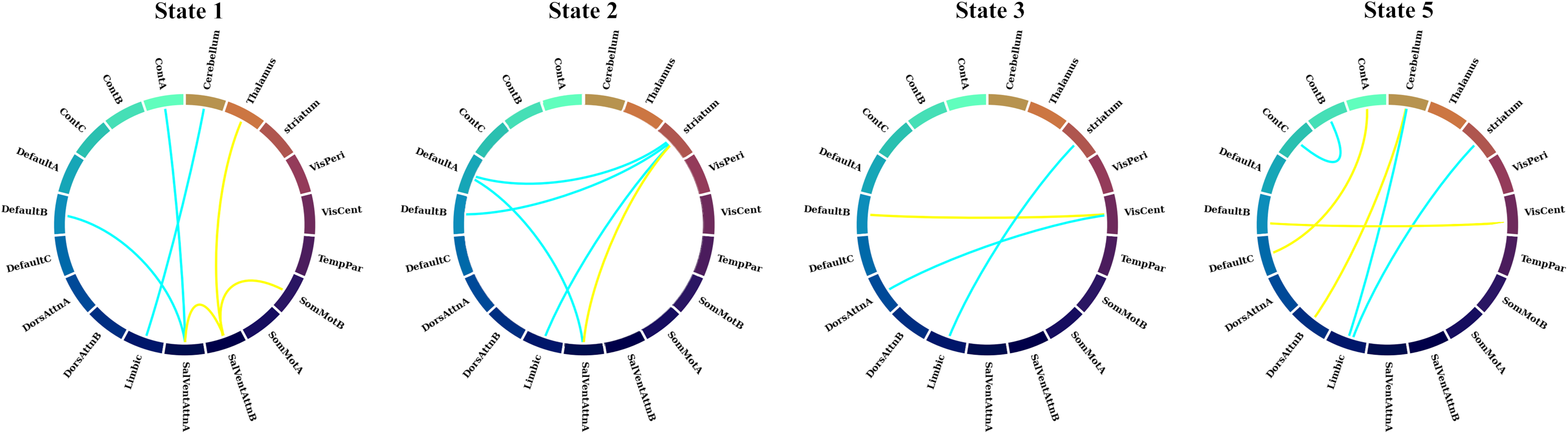
Results of potential pathological features of NT1. Chord diagrams depicting inter-network co-fluctuation differences for NT1 relative to both the SD (acute sleep-deprivation) and healthy control groups, across four dynamic brain states (States 1, 2, 3 and 5). Blue chords indicate network pairs for which NT1 shows significantly higher co-fluctuation than both the SD and control groups. Yellow chords indicate network pairs for which NT1 shows significantly lower co-fluctuation than both the SD and control groups.

### Correlations Between Clinical Measures and Dynamic Functional Connectivity Properties

We further examined whether dFC properties were associated with clinical characteristics in NT1 (Figure 5). Notably, the fraction rate of State 5 was negatively correlated with ESS scores, suggesting that increased occupancy of this state was associated with less subjective sleepiness. In addition, the fraction rate of State 3 showed a significant negative correlation with mean sleep latency from MSLT, whereas the fraction rate of State 2 was positively correlated with mean sleep latency from MSLT, indicating that increased time spent in State 3 may reflect greater arousal instability. Transition probabilities also revealed meaningful associations: transitions from State 4 to State 2 and from State 5 to State 2 were positively correlated with mean sleep latency from MSLT, while transitions from State 5 to State 3 were negatively correlated with mean sleep latency from MSLT.

**Figure 5.**
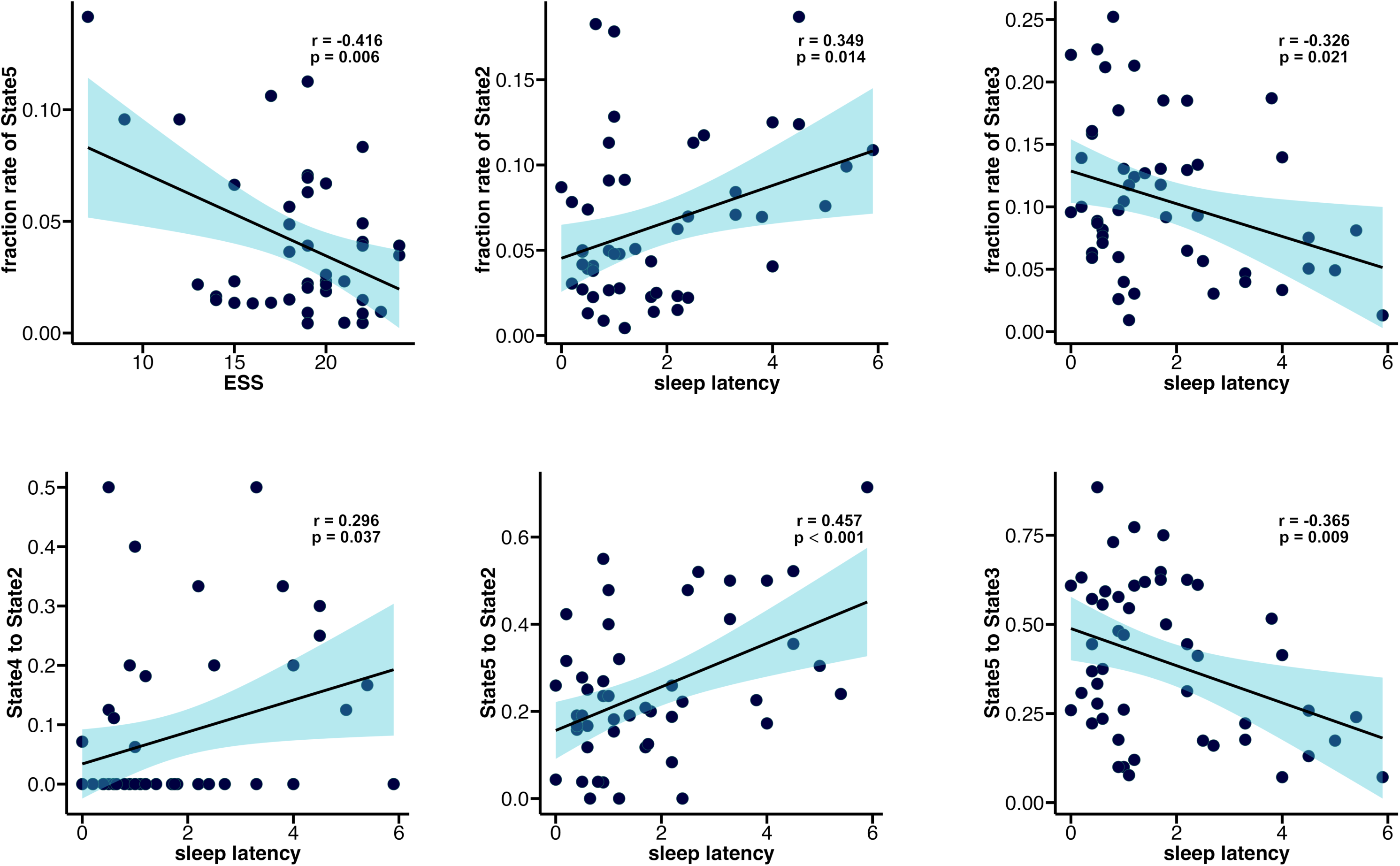
Results of correlations between dynamic functional connectivity temporal properties and clinical characteristics in the NT1 group. Pearson’s correlation test was used, followed by Bonferroni correction.

## Discussion

This study provides novel insights into the brain connectivity dynamics of NT1 by dissociating the effects of hypocretin deficiency from those induced by sleep pressure. To achieve this, we compared patients with NT1 and an SD group, allowing us to identify a pathological brain connectivity pattern (State 3) in NT1. This state was characterized by synchronized co-fluctuations between the salience/ventral attention network (SN/VAN) and the sensorimotor network (SMN), along with antagonistic co-fluctuations of the visual network. This configuration may indicate a shift toward internally driven processing and reduced responsiveness to external sensory input. NT1 showed significantly longer durations in this state (longer mean dwell time and higher fractional rate) compared to both healthy and SD groups, supporting its role as a specific neurophysiological signature of NT1, with temporal properties revealing a distinct pattern of dynamic fluctuations in NT1 and a potential biomarker for clinical severity. Additionally, increased co-fluctuations between striatal and limbic networks were observed in this state, possibly reflecting enhanced dopaminergic engagement to sustain wakefulness. Together, these findings delineate a pathophysiological brain connectivity pattern specific to NT1 and highlight its potential as a functional biomarker for both diagnosis and severity tracking.

By comparing State 3 with State 1 and State 2, respectively, we identified it as a pathological brain state specific to NT1, characterized by disrupted thalamocortical connections and cortical network dysregulation. In detail, State 1 is characterized by functional integration between the SN/VAN and perceptual sensory cortices, with connectivity predominantly anchored in low-level sensory regions, which is in line with the previous study of the effect of sleep deprivation on the hierarchical architectures of the cerebral cortex.^28^ State 2 features coordinated activity between the SN/VAN and the default mode network, reflecting a well-regulated neural pattern that facilitates flexible cognitive switching during wakefulness.^29^ Notably, functional connectivity between the SN/VAN and subcortical structures (i.e., the thalamus and striatum) is shown both in State 1 and 2, whereas this connectivity is absent in State 3 (see Figure 3). Given the crucial role of the SN/VAN as a dynamic hub that integrates thalamic input to coordinate transitions between internally and externally oriented brain states,^30–32^ the absence of such connectivity may impair the SN/VAN’s capacity to regulate large-scale cortical networks in State 3. Moreover, this thalamocortical disconnection is consistent with previous studies reporting reduced thalamocortical connectivity and decreased transmission efficiency in NT1 patients.^7,14,33^ Due to the critical role of thalamocortical circuits for maintaining stable wakefulness,^34^ such a distinct disconnection may represent a key mechanism underlying arousal instability in NT1.

State 3 also exhibits distinct cortical dysregulation. In contrast to the integrative pattern in State 1 and the well-regulated pattern in State 2, State 3 is marked by cortical dysregulation, with antagonistic interactions between the internally oriented network (SN/VAN) and externally oriented sensory processing (visual network and SMN). The shift towards internal processing at the expense of external sensory may reduce the brain’s capacity to maintain awareness and contribute to instability in wakefulness. Such a pattern is consistent with previous findings showing that hypocretin deficiency is associated with impaired cortical sensory integration.^35–37^ Together, these findings highlight State 3 as a pathological brain state in NT1, characterized by disrupted thalamocortical connections and cortical network dysregulation.

NT1 patients exhibited distinct dynamic transition patterns, with an increased tendency to shift into State 3, reflecting abnormal dynamic state regulation that may contribute to the instability of wakefulness in NT1. State 5 represents a baseline resting-state pattern.^38^ NT1 predominantly transitioned from this state to State 3. In contrast, individuals with SD primarily transitioned from State 5 to State 1, and healthy controls transitioned from State 5 to State 2. This divergence in transition pattern suggests that the NT1-specific pattern is likely associated with hypocretin deficiency, rather than sleep pressure. State 3 features synchronized activity between the SN/VAN and SMN networks and antagonism of the VIS network. These features are consistent with evidence that hypocretin deficiency disrupts the coordination of attentional and sensory systems.^39, 40^

In NT1, State 3 was marked by significantly increased co-fluctuations between the striatal and limbic networks compared to both control and SD groups, potentially reflecting compensatory neuromodulatory adaptations aimed at sustaining wakefulness in the context of hypocretin deficiency. This finding is consistent with previous reports of increased activation in these regions in NT1,^8,41^ and may represent a compensatory mechanism for hypocretin deficiency. Given hypocretin’s modulatory role in mesolimbic pathways that regulate reward and arousal,^42^ and its influence on neurotransmitter homeostasis, including dopaminergic,^43, 44^ the increased striatum– limbic co-fluctuations observed in NT1 may reflect a circuit-level attempt to sustain wakefulness through endogenous dopamine signaling. This interpretation is supported by evidence that dopamine contributes to sleep-wake regulation,^45^ and that hypocretin promotes wakefulness partly by facilitating dopamine release via ventral tegmental area activation.^46^ While dopaminergic pathways may account for this change, other neuromodulatory systems are also likely involved, as hypocretin regulates histaminergic, noradrenergic, and acetylcholine transmission.^47–49^

Despite the robust findings, this study has several limitations. First, given that NT1 typically manifests during adolescence,^50^ our findings do not fully capture the dynamic development of this disorder. Second, although comparison with the SD group allowed us to identify NT1-specific connectivity patterns, we were unable to directly assess the relationship between these connectivity alterations and neurotransmitter dynamics. Third, although we retrospectively inspected the functional scans for overt signs of sleep (e.g., closed eyes), the absence of concurrent EEG precluded objective verification of vigilance state during scanning; undetected transitions into light-sleep stages (N1/N2) in the NT1 and SD groups could thus have modulated functional connectivity and biased the group contrasts. A further limitation is the data from two imaging centers, which unavoidably introduced variability in voxel size, coil configuration, and other acquisition parameters. Although we applied ComBat harmonization and implemented a rigorously standardized preprocessing, these differences may still have influenced our functional-connectivity estimates. Future work should employ identical scanner protocols or more advanced harmonization strategies to minimize variance between centers. Finally, we restricted the sample to young adults not only because NT1 is most prevalent in this age group, but also because middle-aged and older individuals often present comorbidities such as hypertension or diabetes. This age restriction might limit the generalizability of our findings; future work should include a wider age range to validate and extend these results. In addition, longitudinal designs and multimodal imaging techniques (e.g., PET to assess dopamine transporter activity and concurrent EEG during fMRI) may provide deeper insights into the association between connectivity alterations and neurotransmitter function in NT1, further elucidating the relationship between NT1 brain connectivity patterns and its pathophysiological mechanisms.

In conclusion, this study reveals a pathological brain connectivity pattern (State 3) specific to NT1, characterized by disrupted thalamocortical interactions, cortical dysregulation, and increased connectivity between striatal and limbic networks. These network-level alterations, likely linked to hypocretin deficiency, correspond closely with clinical severity in NT1. The temporal properties of the pathological state may serve as a neural biomarker for NT1, with potential implications for diagnosis and targeted therapeutic interventions. To clarify the full clinical significance of these findings, future multimodal imaging studies are warranted to further elucidate the complex interactions between connectivity alterations, neurotransmitter systems, and clinical symptomatology in NT1.

### Glossary

CON: executive control network; DMN: default mode network; DAN: dorsal attention network; Limbic: limbic network; SN/VAN: salience/ventral attention network; SMN: somatomotor network; TempPar: temporoparietal network; VIS: visual network; SUB: subcortical network; Cerebellum: cerebellar network; ContA: executive control network A; ContB: executive control network B; ContC: executive control network C; Default A: default mode network A; Default B: default mode network B; Default C: default mode network C; DorsAttnA: dorsal attention network A; DorsAttnB: dorsal attention network B; Sal/VentAttnA: salience/ventral attention network A; Sal/VentAttnB: salience/ventral attention network B; SomMotA: somatomotor network A; SomMotB: somatomotor network B; VisCent: visual center network; VisPeri: visual peripheral network.

## Acknowledgments

The authors would like to express their sincere gratitude to Prof. Ze Wang, University of Maryland School of Medicine, for his valuable advice and assistance with ComBat correction for data harmonization.

## Data Availability

The dataset and codes used for this study will be made available by the corresponding author on request.

## Author Contributions

Wenwei Zhu: investigation, formal analysis, writing – original draft, writing – review and editing. Fulong Xiao: investigation, formal analysis, funding acquisition, writing – review and editing. Mengmeng Wang: data curation. Xiaosong Dong: software, resources, methodology. Fang Han: conceptualization, funding acquisition, resources; supervision. Ning Ma: conceptualization, funding acquisition, resources; writing-review and editing, supervision. All authors had full access to all data in the study and took responsibility for the integrity of the data and the accuracy of the data analysis. All authors agreed on the journal to which the article will be submitted, reviewed and agreed on all versions of the article before submission, during revision, the final version accepted for publication, and any significant changes introduced at the proofing stage, agreed to take responsibility and be accountable for the contents of the article.

