## Supplemental Methods; Supplemental Figure 1;Supplemental Table 1-7 for "Dynamic Functional Connectivity States in Narcolepsy Type 1: Distinct Patterns from Acute Sleep Deprivation and Associations with Clinical Measures of Sleepiness"

**This PDF file includes:**

Supplemental Methods

Supplemental Figure 1

Supplemental Tables 1 to 7

### *I. Supplemental Methods*

#### The diagnosis of NT1 was based on the ICSD-3

The diagnosis of NT1 was based on the ICSD-3: excessive daytime sleepiness lasting at least 3 months, typical cataplexy, with mean sleep latency ≤8 min and 2 or more sleep-onset REM periods (SOREMPs) on the multiple sleep latency test (MSLT) or during the previous night polysomnography (PSG), or lower levels of orexin (≤110 pg/mL) in cerebral spinal fluid. NT1 patients with obstructive sleep apnea and hypopnea syndrome (AHI ≥5), cognitive dysfunction (measured by Mini-Mental State Examination, MMSE < 27), mental disorders (assessed by psychiatrists based on clinical manifestations and questionnaires), substance/alcohol abuse or other serious disease were excluded.

The exclusion criteria for both patients with NT1 and the two healthy groups were as follows: 1) alcohol, drug, or substance abuse; 2) congenital or inherited diseases; 3) other sleep disorders, such as obstructive sleep apnea or insomnia; 4) neurological diseases or structural brain lesions based on imaging; 5) diabetes, chronic pulmonary/respiratory disease, or heart disease; 6) psychiatric comorbidities (e.g., depression, anxiety); or 7) MRI contraindications, such as claustrophobia or metal implants.

#### Sleep and Circadian Assessments

The sleep quality in the healthy controls and SD groups was assessed by sleep physicians using sleep questionnaires, such as the Epworth Sleepiness Scale (ESS), Pittsburgh Sleep Quality Index (PSQI), and the STOP-Bang questionnaire to exclude excessive daytime sleepiness, insomnia, and sleep apnea. Besides, the Morningness – Eveningness Questionnaire (MEQ) was used to assess circadian preference in NT1 patients and healthy controls and SD groups, excluding participants whose scores indicated divergent chronotypes. None of the participants in the healthy control and SD groups had any history of psychiatric or neurological disorders.

#### Polysomnography and MSLT Evaluation

All patients were required to abstain from coffee or alcoholic drinks for one day before participation in this study. A full nocturnal polysomnography was recorded on a Respironics LE-Series Physiological Monitoring System (Alice 6 LE, Philips, Murrysville, FL) from 10:00 p.m. to 6:00 a.m. In accordance with guidelines established by the American Academy of Sleep Medicine (AASM), the standard electroencephalogram (EEG) was recorded from the frontal, central, and occipital regions (F4/M1, C4/M1, and O2/M1, and back-up F3/M2, C3/M2, and O1/M2), chin electromyograms (EMG from 3chin electrodes and the middle of the right anterior tibialis), the electrooculogram (EOG located in the cornea and retina), and electrocardiogram. Furthermore, the following parameters were recorded: oral and nasal airflow, snoring, chest and abdominal breathing, oxygen saturation, and body position, along with total sleep time, sleep latency, sleep efficiency, arousal, and respiratory events. According to the AASM manual, obstructive apnea was defined as a reduction in airflow of at least 90% lasting at least 10 seconds and associated with persistent respiratory effort. Hypopnea was defined as a reduction in airflow of at least 30% lasting at least 10 seconds and accompanied by a 4% or greater oxygen desaturation.[1] The Apnea-Hypopnea Index (AHI) was calculated as the average of the total number of apnea and hypopnea events experienced per hour of sleep. All patients were requested to complete a clinical MSLT on the day following the nocturnal polysomnography. The AASM task-force-approved modification was followed,[2] with naps scheduled at 2-hour intervals beginning 2 hours after the initial morning awakening. If sleep did not occur within 20 minutes, the nap trial was terminated and sleep latency was recorded as 20 minutes. If sleep occurred within 20 minutes, the onset of sleep was defined as the time from lights-out to the first epoch of sleep, including stage 1. To assess the presence of REM sleep, the test continued for 15 minutes after sleep onset. The latency to REM sleep was recorded if it occurred. Subsequently, the mean sleep latency and REM sleep latency from the 5 naps in MSLT were calculated.

#### Experimental protocol of Acute Sleep Deprivation

Besides the exclusion criteria mentioned before, the healthy participants who suffered Acute sleep deprivation (ASD) should also fit 1) good sleep habits (7–9 h of total sleep time with sleep onset no later than 1:00 AM and wake time between 7:00 and 9:00 AM) for at least a month; and 2) no trans-meridian travel, shift work, or irregular sleep-wake routines in the 60 days before the experiment. All participants were kept from any caffeine or medicine intake for 72 hours before ASD. In the ASD session, participants reached the laboratory at 7:00 p.m. They had to stay awake all night under the supervision of trained personnel, which resulted in approximately 24 hours of total sleep deprivation. During this session, activities were limited to reading, writing, watching low-arousal movies, playing low-arousal games, and short walking. All activities were continuously monitored by trained personnel through surveillance cameras throughout the sleep-deprived night. Oral warnings were given if participants closed their eyes for a while or lay on the desk to ensure them stayed awake.

#### Imaging Data Acquisition

The MRI data of NT1 patients and healthy control participants were obtained on a 3T scanner (Siemens, Skyra, Germany) using an 8-channel brain phased-array coil. Foam pads were used to minimize head motion, and headphones were used to reduce scanner noise. Resting BOLD MRI scans were obtained with gradient-echo planar imaging (TR = 2030 ms, TE = 30 ms, slice = 33, FA = 90°, FOV = 224mm × 224mm, matrix = 64 × 64, voxel size = 3.5 × 3.5 × 3.5). After the BOLD MRI scan, a high-resolution T1-weighted structural image was acquired with the following parameters: TR = 1900 ms, TE = 2.55 ms, FA = 9°, FOV = 240 mm × 240 mm, thickness = 1 mm. All patients were examined by MRI immediately following the clinical MSLT. All patients and healthy control participants were asked to resist sleeping to remain fully awake, not to move, and to keep their eyes open during the whole MRI scan,[3, 4] supervised clinically and by video during the whole process. In addition, emotional triggering factors were avoided during the whole process to prevent a cataplexy attack.

MRI examination for ASD participants was performed the morning after ASD. All MRI imaging data were obtained on a 3T scanner (Siemens, Erlangen, Germany) using a 12-channel brain phased-array coil. Foam pads were used to minimize head motion, and headphones were used to reduce scanner noise. Resting BOLD MRI scans were obtained with T2*-weighted echo-planar imaging (TR = 2,000 ms, TE = 24 ms, slice = 30, FA = 90°, FOV = 220mm × 220 mm, matrix = 64 × 64, voxel size = 3.4 × 3.4 × 4). After the BOLD MRI scan, a high-resolution T1-weighted structural image was acquired using the magnetization prepared rapid gradient-echo (MPRAGE) sequence with the following parameters: TR = 2,300 ms, TE = 3.24 ms, FA = 9°, FOV = 256mm × 256 mm, thickness = 1 mm. All acute sleep-deprived participants were asked to remain fully awake, remain still, and keep their eyes open throughout the entire process.

#### Preprocessing of Functional Imaging Data

A total of 240 functional volumes were acquired in the resting BOLD MRI scans. The first ten functional volume images of each participant's dataset were discarded, then the remaining fMRI data were corrected for slice timing and realigned for motion correction. Participants with head motion exceeding 0.5 mm in translation and 1 in rotation were rejected. Anatomical and functional images were first manually reoriented to the anterior commissure, and structural images were co-registered to the functional images for each participant using a linear transformation. Then, the co-registered functional images were normalized to the standard Montreal Neurological Institute space template with a resampling voxel size of 3 mm × 3 mm × 3 mm. The normalized functional images were smoothed using a Gaussian filter 4 mm full width at half maximum. All smoothed images were filtered using a typical temporal bandpass (0.01 Hz–0.1 Hz) to reduce low-frequency drift and physiological high-frequency respiratory and cardiac noise. Linear trends were removed within each time series. The covariates were regressed out from the time series of every voxel, including the white matter signal, cerebrospinal fluid signal, 24 motion parameters,[5] and the global signal.

#### Edge (Co-fluctuation) time series

Functional brain networks are constructed by estimating the statistical dependency between fMRI BOLD activity of brain regions. The magnitude of these dependencies reflects the strength of the functional connection between brain regions. One of the most common measures to estimate the dependency between brain regions is the Pearson correlation coefficient. If there are *T* time points, *i* and *j* are two nodes, the overall procedure for calculating the Pearson coefficient is as follows: Let $x_i = [x_i(1), \cdots, x_i(T)]$ and $x_j = [x_j(1), \cdots, x_j(T)]$ be the time series recorded from voxels or parcels $i$ and $j$, respectively. We can calculate the correlation of $i$ and $j$ by first z-scoring each time series, such that $z_i = \frac{x_i - \mu_i}{\sigma_i}$, where $\mu_i = \frac{1}{T}\sum_t x_i(t)$ and $\sigma_i = \frac{1}{T-1}\sum_t x_i(t) - \mu_i$ are the time averaged mean and standard deviation. Then, the correlation of $i$ with $j$ can be calculated as $r_{ij} = \frac{1}{T-1}\sum_t [z_i(t) \times z_j(t)]$. Repeating this procedure for all pairs of parcels results in a node-by-node correlation matrix, i.e., an estimate of FC. If there are N nodes, this matrix has dimensions $N \times N$. To estimate edge-centric networks, we modify the above approach such that we only calculate the element-wise product of two time-series and remove the step for calculating the mean. This operation would result in a vector of length $T$ whose elements encode the moment-by-moment co-fluctuation magnitude of parcels $i$ and $j$. More specifically, the positive values in the vector reflect the simultaneous increase or decrease in the activity of parcels $i$ and $j$, while negative values reflect the opposite direction (one increasing while the other decreasing and vice versa) of the magnitude of their activity. Similarly, if either $i$ or $j$ increased or decreased while the activity of the other was close to the baseline, the corresponding entry would be close to zero. An analogous vector can easily be calculated for every pair of parcels (network nodes), resulting in a set of edge time series. With $N$ parcels, this results in $\frac{N \times (N-1)}{2}$ pairs, each of length $T$ (a matrix of $\frac{N \times (N-1)}{2} \times T$). Each column represents the functional connectivity at every time point.

### ComBat Correction for Data Batch Effects

The ComBat model was introduced in the context of gene expression analysis by Johnson, Li and Rabinovic [6] as an improvement of location/scale models for studies with a small sample size. Here, we reformulate the ComBat model in the context of resting images.ComBat is based on the empirical Bayes framework that adjusts for batch effects while preserving the biological signal in the data. It models batch effects as systematic shifts and then adjusts for these differences across batches, ensuring that the adjusted data more accurately reflects the true underlying patterns of brain activity. We assume that the data come from $m$ imaging sites, containing each $n_i$ scans for $i$ = 1, 2, … , $m$. For voxel $v$ = 1, 2, … , $p$, let $y_{ijv}$ represent the FA measure for the scan $j$ at site $i$. After some standardization discussed in Johnson et al. (2007), ComBat posits the following location and scale (L/S) adjustment model: $y_{ijv} = \alpha_v + X_{ij}\beta_v + \gamma_{iv} + \delta_{iv}\varepsilon_{ijv}$ , where $\alpha_v$ is the overall FA measure for voxel $v$, $X$ is a design matrix for the covariates of interest (e.g. sex, age), and $\beta_v$ is the voxel-specie vector of regression coefficients corresponding to $X$. We further assume that the error terms $\varepsilon_{ijv}$ follow a normal distribution with mean zero and variance $\delta_v^2$ . The terms $\gamma_{iv}$ and $\delta_{iv}$ represent the additive and multiplicative site effects of site $i$ for voxel $v$, respectively.

ComBat uses an empirical Bayes (EB) framework to improve the variance of the parameter estimates $\gamma_{iv}$ and $\delta_{iv}$. It estimates an empirical statistical distribution for each of those parameters by assuming that all voxels share the same common distribution. In that sense, information from all voxels is used to inform the statistical properties of the site effects. More specifically, the site-effect parameters are assumed to have the parametric prior distributions: $\gamma_{iv} \sim N(\gamma_i, \tau_i^2)$ and $\delta_{iv}^2 \sim Inverse\ Gamma(\lambda_i \vartheta_i)$. The hyperparameters $\gamma_i$; $\tau_i^2$; $\lambda_i; \vartheta_i$ are estimated empirically from the data as described in Johnson, Li and Rabinovic [6]. We note that the sva package also offers the option to posit non-parametric priors for more flexibility, at the cost of increasing computational time. The ComBat estimates $\gamma_{iv}$ and $\delta_{iv}$ of the site effect parameters are computed using conditional posterior means, the final ComBat-harmonized FA values are defined as: $y_{ijv}^{\text{ComBat}} = \frac{y_{ijv} - \hat{\alpha}_v - \mathbf{X}_{ij}\hat{\boldsymbol{\beta}}_v - \gamma_{iv}^*}{\delta_{iv}^*} + \hat{\alpha}_v + \mathbf{X}_{ij}\hat{\boldsymbol{\beta}}_v$.

#### Clustering Analysis of ETS

We used the L1 distance (Manhattan distance) function to estimate the similarity between window functional connectivity matrices (window size = 1TR), as it has been demonstrated to be an effective measure for high-dimensional data. More specifically, time points were clustered based on the similarity of whole-brain co-fluctuation patterns at each time point. For each participant, we obtained a clustered time series ($1 \times T$), where each element represents a cluster index (i.e., brain state) at that given time point. After clustering ETS, we quantified the number of transitions between states over time.

### *II. Supplemental Figure*

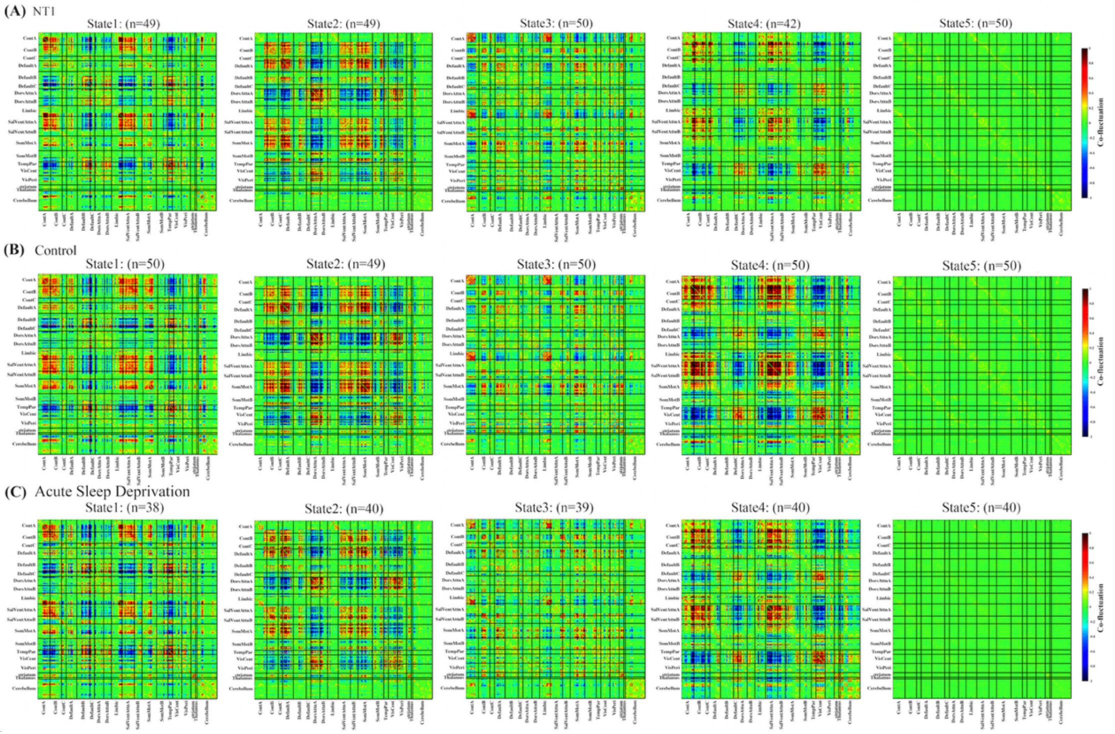


**Supplemental Figure 1. Group-specific cluster centroid for each state. (A)** Group-specific cluster centroid for each state, averaged across the subject-specific median cluster centroid in NT1. **(B)** Group-specific cluster centroid for each state, averaged across the subject-specific median cluster centroid in Control. **(C)** Group-specific cluster centroid for each state, averaged across subject-specific median cluster centroid in acute sleep deprivation.

### *II. Supplemental Tables*

**Supplemental Table 1.** Group differences in mean dwell time across states

| State | NT1 M(SD) | Control M(SD) | SD M(SD) | *F* | *P* | Post-Hoc | Post-Hoc-*p* |
|---|---|---|---|---|---|---|---|
| State1 | 1.83(0.44) | 1.67(0.46) | 1.83(0.49) | F (2, 135) = 1.82 | 0.16 | NT1 *vs.* Control | 0.29 |
| | | | | | | NT1 *vs.* ASD | 0.99 |
| | | | | | | Control *vs.* ASD | 0.34 |
| State2 | 1.94(0.37) | 2.04(0.28) | 1.75(0.49) | F (2, 134) = 7.88 | **< 0.001** | NT1 *vs.* Control | 0.65 |
| | | | | | | NT1 *vs.* ASD | **0.02** |
| | | | | | | Control *vs.* ASD | **<0.001** |
| State3 | 2.13(0.46) | 1.72(0.41) | 1.58(0.33) | F (2, 136) = 22.2 | **< 0.001** | NT1 *vs.* Control | **<0.001** |
| | | | | | | NT1 *vs.* ASD | **<0.001** |
| | | | | | | Control *vs.* ASD | 0.38 |
| State4 | 1.73(0.46) | 1.91(0.41) | 1.72(0.47) | F (2, 128) = 2.56 | 0.08 | NT1 *vs.* Control | 0.19 |
| | | | | | | NT1 *vs.* ASD | 0.96 |
| | | | | | | Control *vs.* ASD | 0.15 |
| State5 | 7.52(2.11) | 6.94(2.55) | 7.28(1.97) | F (2, 137) = 0.84 | 0.43 | NT1 *vs.* Control | 0.59 |
| | | | | | | NT1 *vs.* ASD | 0.86 |
| | | | | | | Control *vs.* ASD | 0.95 |

**Note:** This table presents the mean dwell time (*M* ± *SD*) for each state across the NT1, healthy control, and Acute sleep deprivation (SD) groups. A one-way ANOVA was performed to assess group differences for each state. Significant effects were followed by Bonferroni-corrected post hoc comparisons. Statistically significant results are marked in bold.

**Supplemental Table 2.** Group differences in fraction rate across states

| State | NT1 M(SD) | Control M(SD) | SD M(SD) | *F* | *P* | Post-Hoc | Post-Hoc-*p* |
|---|---|---|---|---|---|---|---|
| State1 | 0.05(0.03) | 0.05(0.02) | 0.08(0.05) | F (2, 135) = 7.62 | **< 0.001** | NT1 *vs.* Control | 0.33 |
| | | | | | | NT1 *vs.* ASD | **0.008** |
| | | | | | | Control *vs.* ASD | **< 0.001** |
| State2 | 0.06(0.04) | 0.11(0.04) | 0.05(0.03) | F (2, 134) = 29.01 | **< 0.001** | NT1 *vs.* Control | **< 0.001** |
| | | | | | | NT1 *vs.* ASD | 0.25 |
| | | | | | | Control *vs.* ASD | **< 0.001** |
| State3 | 0.11(0.06) | 0.05(0.04) | 0.05(0.03) | F (2, 136) = 18.43 | **< 0.001** | NT1 *vs.* Control | **< 0.001** |
| | | | | | | NT1 *vs.* ASD | **< 0.001** |
| | | | | | | Control *vs.* ASD | 0.65 |
| State4 | 0.04(0.03) | 0.06(0.05) | 0.06(0.04) | F (2, 128) = 2.84 | 0.06 | NT1 *vs.* Control | 0.1 |
| | | | | | | NT1 *vs.* ASD | 0.62 |
| | | | | | | Control *vs.* ASD | 0.15 |
| State5 | 0.74(0.06) | 0.73(0.07) | 0.76(0.05) | F (2, 137) = 2.99 | 0.06 | NT1 *vs.* Control | 0.58 |
| | | | | | | NT1 *vs.* ASD | 0.68 |
| | | | | | | Control *vs.* ASD | 0.06 |

**Note:** This table summarizes the fraction rate (*M* ± *SD*) for each state across the NT1, healthy control, and Acute sleep deprivation (SD) groups. The fraction rate reflects the proportion of total time spent in each state. A one-way ANOVA was performed to assess group differences for each state. Significant effects were followed by Bonferroni-corrected post hoc comparisons. Statistically significant results are marked in bold.

**Supplemental Table 3.** Transition probabilities between states across NT1, control, and ASD groups

| State | NT | Control | SD | *F* | *P* | Post-Hoc | Post-Hoc-*p* |
|---|---|---|---|---|---|---|---|
| | | | | | | NT1 *vs.* Control | 0.78 |
| S1 to S2 | 0.06(0.11) | 0.13(0.19) | 0.06(0.10) | F (2, 137) = 3.32 | 0.56 | NT1 *vs.* ASD | 0.95 |
| | | | | | | Control *vs.* ASD | 0.96 |
| | | | | | | NT1 *vs.* Control | 0.31 |
| S1 to S3 | 0.12(0.15) | 0.07(0.12) | 0.08(0.18) | F (2, 137) = 1.54 | 0.22 | NT1 *vs.* ASD | 0.57 |
| | | | | | | Control *vs.* ASD | 0.82 |
| | | | | | | NT1 *vs.* Control | 0.29 |
| S1 to S4 | 0.03(0.08) | 0.06(0.12) | 0.04(0.07) | F (2, 137) = 1.50 | 0.23 | NT1 *vs.* ASD | 0.62 |
| | | | | | | Control *vs.* ASD | 0.73 |
| | | | | | | NT1 *vs.* Control | 0.91 |
| S1 to S5 | 0.77(0.22) | 0.72(0.24) | 0.82(0.21) | F (2, 137) = 2.25 | 0.11 | NT1 *vs.* ASD | 0.76 |
| | | | | | | Control *vs.* ASD | 0.11 |
| | | | | | | NT1 *vs.* Control | 0.92 |
| S2 to S1 | 0.05(0.09) | 0.06(0.06) | 0.12(0.16) | F (2, 137) = 5.35 | **0.006** | NT1 *vs.* ASD | **0.009** |
| | | | | | | Control *vs.* ASD | 0.52 |
| | | | | | | NT1 *vs.* Control | 0.086 |
| S2 to S3 | 0.13(0.22) | 0.06(0.09) | 0.05(0.10) | F (2, 137) = 3.92 | **0.02** | NT1 *vs.* ASD | **0.03** |
| | | | | | | Control *vs.* ASD | 0.91 |
| | | | | | | NT1 *vs.* Control | 0.84 |
| S2 to S4 | 0.06(0.11) | 0.07(0.12) | 0.08(0.14) | F (2, 137) = 0.28 | 0.75 | NT1 *vs.* ASD | 0.93 |
| | | | | | | Control *vs.* ASD | 0.88 |

*(continued on next page)*

| State | NT1 | Control | SD | *F* | *P* | Post-Hoc | Post-Hoc-*p* |
|---|---|---|---|---|---|---|---|
| | | | | | | NT1 *vs.* Control | 0.37 |
| S2 to S5 | 0.74(0.25) | 0.81(0.15) | 0.70(0.26) | F (2, 137) = 2.620 | 0.07 | NT1 *vs.* ASD | 0.74 |
| | | | | | | Control *vs.* ASD | 0.08 |
| | | | | | | NT1 *vs.* Control | 0.15 |
| S3 to S1 | 0.06(0.09) | 0.04(0.11) | 0.11(0.13) | F (2, 137) = 4.79 | **0.01** | NT1 *vs.* ASD | 0.12 |
| | | | | | | Control *vs.* ASD | **0.02** |
| | | | | | | NT1 *vs.* Control | **< 0.001** |
| S3 to S2 | 0.06(0.01) | 0.21(0.25) | 0.09(0.18) | F (2, 137) = 8.03 | **< 0.001** | NT1 *vs.* ASD | 0.96 |
| | | | | | | Control *vs.* ASD | **0.02** |
| | | | | | | NT1 *vs.* Control | 0.93 |
| S3 to S4 | 0.06(0.12) | 0.04(0.08) | 0.09(0.14) | F (2, 137) = 2.13 | 0.12 | NT1 *vs.* ASD | 0.42 |
| | | | | | | Control *vs.* ASD | 0.14 |
| | | | | | | NT1 *vs.* Control | **0.04** |
| S3 to S5 | 0.82(0.14) | 0.72(0.26) | 0.68(0.24) | F (2, 137) = 5.37 | **0.006** | NT1 *vs.* ASD | **0.008** |
| | | | | | | Control *vs.* ASD | 0.58 |
| | | | | | | NT1 *vs.* Control | 0.28 |
| S4 to S1 | 0.16(0.26) | 0.093(0.18) | 0.09(0.13) | F (2, 137) = 1.98 | 0.14 | NT1 *vs.* ASD | 0.26 |
| | | | | | | Control *vs.* ASD | 0.89 |
| | | | | | | NT1 *vs.* Control | 0.39 |
| S4 to S2 | 0.08(0.14) | 0.13(0.19) | 0.10(0.19) | F (2, 137) = 1.21 | 0.30 | NT1 *vs.* ASD | 0.84 |
| | | | | | | Control *vs.* ASD | 0.91 |

*(continued on next page)*

| State | NT1 | Control | SD | *F* | *P* | Post-Hoc | Post-Hoc-*p* |
|---|---|---|---|---|---|---|---|
| | | | | | | NT1 *vs.* Control | 0.87 |
| S4 to S3 | 0.10(0.19) | 0.08(0.13) | 0.08(0.12) | F (2, 137) = 0.34 | 0.72 | NT1 *vs.* ASD | 0.95 |
| | | | | | | Control *vs.* ASD | 0.82 |
| | | | | | | NT1 *vs.* Control | **0.002** |
| S4 to S5 | 0.49(0.36) | 0.69(0.24) | 0.71(0.24) | F (2, 137) = 8.13 | **< 0.001** | NT1 *vs.* ASD | **0.002** |
| | | | | | | Control *vs.* ASD | 0.52 |
| | | | | | | NT1 *vs.* Control | 0.32 |
| S5 to S1 | 0.22(0.13) | 0.21(0.12) | 0.32(0.18) | F (2, 137) = 7.45 | **< 0.001** | NT1 *vs.* ASD | **0.006** |
| | | | | | | Control *vs.* ASD | **0.001** |
| | | | | | | NT1 *vs.* Control | **< 0.001** |
| S5 to S2 | 0.24(0.17) | 0.38(0.14) | 0.18(0.13) | F (2, 137) = 22.25 | **< 0.001** | NT1 *vs.* ASD | 0.14 |
| | | | | | | Control *vs.* ASD | **< 0.001** |
| | | | | | | NT1 *vs.* Control | **< 0.001** |
| S5 to S3 | 0.39(0.22) | 0.21(0.12) | 0.25(0.14) | F (2, 137) = 16.73 | **< 0.001** | NT1 *vs.* ASD | **< 0.001** |
| | | | | | | Control *vs.* ASD | 0.69 |
| | | | | | | NT1 *vs.* Control | 0.08 |
| S5 to S4 | 0.14(0.12) | 0.20(0.16) | 0.25(0.15) | F (2, 137) = 6.76 | 0.002 | NT1 *vs.* ASD | **< 0.001** |
| | | | | | | Control *vs.* ASD | 0.43 |

**Note:** This table summarizes the transition probabilities between each state for the NT1, control, and Acute sleep deprivation (SD) groups. Values are given as mean (Standard Deviation). For each state-to-state transition (e.g., S1 to S2), mean values and standard deviations are reported. Group differences were assessed using one-way ANOVA, and significant effects were followed up with post-hoc pairwise comparisons. Statistically significant p-values are highlighted in bold.

**Supplemental Table 4.** NT1-specific alterations in co-fluctuation strength during State 1

| | co-fluctuation | *F* | *P* | Post-Hoc-*p* (NT1 *vs.* Control) | Post-Hoc-*p* (NT1 *vs.* SD) | Post-Hoc-*p* (Control vs. SD) |
|---|---|---|---|---|---|---|
| NT1 > Control & NT1 > SD | SalVentAttnA — ContA | F (2, 135) = 4.05 | **< 0.001** | **0.02** | **0.01** | 0.55 |
| | SalVentAttnB — DefaultB | F (2, 135) = 5.53 | **0.006** | **0.002** | **0.04** | 0.35 |
| | Cerebellum — Limbic | F (2, 135) = 5.72 | **< 0.001** | **0.02** | **< 0.001** | 0.27 |
| NT1 < Control & NT1 < SD | SalVentAttnA — SalVentAttnB | F (2, 135) = 4.55 | **0.02** | **0.04** | **0.03** | 0.25 |
| | SalVentAttnB — SomMotB | F (2, 135) = 5.99 | **< 0.001** | **0.03** | **0.006** | 0.15 |
| | SalVentAttnB — Thalamus | F (2, 135) = 5.16 | **0.002** | **0.03** | **0.006** | 0.16 |

**Note:** This table summarizes the co-fluctuations that are uniquely altered in NT1 patients during State 1, as determined by pairwise comparisons with both healthy controls and Acute sleep deprivation (SD) participants. Network pairs where NT1 showed significantly greater co-fluctuation than both other groups are listed under "NT1 > Control & NT1 > SD", while pairs with significantly reduced co-fluctuation in NT1 are listed under "NT1 < Control & NT1 < SD". Statistical comparisons were conducted using ANOVA followed by post-hoc tests. Only connections showing significant differences ($p < 0.05$) in both comparisons (NT1 vs. Control and NT1 vs. SD) are included. Statistically significant p-values are highlighted in bold.

**Supplemental Table 5.** NT1-specific alterations in co-fluctuation strength during State 2

| | co-fluctuation | *F* | *P* | Post-Hoc-*p* (NT1 *vs.* Control) | Post-Hoc-*p* (NT1 *vs.* SD) | Post-Hoc-*p* (Control vs. SD) |
|---|---|---|---|---|---|---|
| NT1 > Control & NT1 > SD | SalVentAttnA — DefaultA | F (2, 134) = 11.38 | **< 0.001** | **< 0.001** | **0.03** | **0.04** |
| | Striatum — DefaultA | F (2, 134) = 4.26 | **0.003** | **0.001** | **0.04** | 0.97 |
| | Striatum — DefaultB | F (2, 134) = 4.89 | **0.02** | **0.03** | **0.007** | 0.14 |
| | Striatum — Limbic | F (2, 134) = 4.39 | **0.03** | **0.02** | **0.01** | 0.23 |
| NT1 < Control & NT1 < SD | Striatum —SalVentAttnA | F (2, 134) = 4.22 | **0.006** | **0.003** | **0.02** | 0.91 |

**Note:** This table presents the co-fluctuations that are uniquely altered in NT1 patients during State 2, based on comparisons with both healthy controls and Acute sleep deprivation (SD) participants. The upper section lists network pairs where NT1 showed significantly greater co-fluctuation strength than both the control and SD groups ("NT1 > Control & NT1 > SD"), while the lower section lists pairs with significantly weaker co-fluctuation in NT1 ("NT1 < Control & NT1 < SD"). All reported effects met the significance threshold in both comparisons. Statistically significant p-values are highlighted in bold.

**Supplemental Table 6.** NT1-specific alterations in co-fluctuation strength during State 3

| | co-fluctuation | *F* | *P* | Post-Hoc-*p* (NT1 *vs.* Control) | Post-Hoc-*p* (NT1 *vs.* SD) | Post-Hoc-*p* (Control vs. SD) |
|---|---|---|---|---|---|---|
| NT1 > Control & NT1 > SD | Striatum — Limbic | F (2, 136) = 19.85 | **< 0.001** | **0.003** | **< 0.001** | **0.001** |
| | VisCent — DorsAttnA | F (2, 136) = 5.43 | **0.005** | **0.04** | **0.002** | 0.21 |
| NT1 < Control & NT1 < SD | VisCent — DefaultB | F (2, 136) = 5.01 | **< 0.001** | **0.03** | **< 0.001** | 0.06 |

**Note:** This table summarizes the co-fluctuations that are uniquely altered in NT1 patients during State 3, as determined by pairwise comparisons with both healthy controls and Acute sleep deprivation (SD) participants. Network pairs where NT1 showed significantly greater co-fluctuation than both other groups are listed under "NT1 > Control & NT1 > SD", while pairs with significantly reduced co-fluctuation in NT1 are listed under "NT1 < Control & NT1 < SD". Statistical comparisons were conducted using ANOVA followed by post-hoc tests. Only connections showing significant differences ($p < 0.05$) in both comparisons (NT1 vs. Control and NT1 vs. SD) are included. Statistically significant p-values are highlighted in bold.

**Supplemental Table 7.** NT1-specific alterations in co-fluctuation strength during State 5

| | co-fluctuation | *F* | *P* | Post-Hoc-*p* (NT1 *vs.* Control) | Post-Hoc-*p* (NT1 *vs.* SD) | Post-Hoc-*p* (Control vs. SD) |
|---|---|---|---|---|---|---|
| NT1 > Control & NT1 > SD | ContC — ContB | F (2, 137) = 8.52 | **< 0.001** | **< 0.001** | **0.002** | 0.47 |
| | Striatum — Limbic | F (2, 137) = 24.51 | **< 0.001** | **0.002** | **< 0.001** | **< 0.001** |
| | Cerebellum — Limbic | F (2, 137) = 5.57 | **< 0.001** | **0.001** | **0.002** | 0.17 |
| NT1 < Control & NT1 < SD | DefaultC — ContA | F (2, 137) = 6.25 | **0.003** | **0.02** | **< 0.001** | 0.38 |
| | VisCent — DefaultB | F (2, 137) = 5.35 | **0.004** | **0.005** | **0.006** | 0.99 |
| | Cerebellum — DorsAttnB | F (2, 137) = 6.18 | **0.002** | **0.001** | **0.02** | 0.37 |

**Note:** This table summarizes the co-fluctuations that are uniquely altered in NT1 patients during State 3, as determined by pairwise comparisons with both healthy controls and Acute sleep deprivation (SD) participants. Network pairs where NT1 showed significantly greater co-fluctuation than both other groups are listed under "NT1 > Control & NT1 > SD", while pairs with significantly reduced co-fluctuation in NT1 are listed under "NT1 < Control & NT1 < SD". Statistical comparisons were conducted using ANOVA followed by post-hoc tests. Only connections showing significant differences ($p < 0.05$) in both comparisons (NT1 vs. Control and NT1 vs. SD) are included. Statistically significant p-values are highlighted in bold.
